# Elevated symptoms of depression and anxiety among family members of critically ill COVID-19 patients - An observational study of five cohorts across four countries

**DOI:** 10.1101/2023.02.28.23286559

**Authors:** Anikó Lovik, Juan González-Hijón, Asle Hoffart, Chloe Fawns-Ritchie, Ingibjörg Magnúsdóttir, Li Lu, Anna Bára Unnarsdóttir, Anna K. Kähler, Archie Campbell, Arna Hauksdóttir, Charilaos Chourpiliadis, Daniel L. McCartney, Edda Björk Thordardóttir, Emily E. Joyce, Emma M. Frans, Jóhanna Jakobsdóttir, Lill Trogstad, Ole A. Andreassen, Per Magnus, Sverre Urnes Johnson, Patrick F. Sullivan, Thor Aspelund, David J. Porteous, Helga Ask, Omid V. Ebrahimi, Unnur Anna Valdimarsdóttir, Fang Fang

**Author notes:** Equal contribution (Anikó Lovik, Juan González-Hijón, Asle Hoffart, Chloe Fawns-Ritchie, Ingibjörg Magnúsdóttir, Li Lu, Anna Bára Unnarsdóttir). Equal contribution (David J. Porteous, Helga Ask, Omid V. Ebrahimi, Unnur Anna Valdimarsdóttir, Fang Fang).

## Abstract

**Background:** Little is known regarding the mental health impact of having a significant person (family member and/or close friend) with COVID-19 of different severity.

**Methods:** The study included five prospective cohorts from four countries (Iceland, Norway, Sweden, and the UK) with self-reported data on COVID-19 and symptoms of depression and anxiety during March 2020-March 2022. We calculated the prevalence ratio (PR) of depression and anxiety in relation to having a significant person with COVID-19 and performed a longitudinal analysis in the Swedish cohort to describe the temporal patterns of the results.

**Results:** 162,237 and 168,783 individuals were included in the analysis of depression and anxiety, respectively, of whom 24,718 and 27,003 reported a significant person with COVID-19. Overall, the PR was 1.07 (95% CI: 1.05-1.10) for depression and 1.08 (95% CI: 1.03-1.13) for anxiety among significant others of COVID-19 patients. The respective PRs for depression and anxiety were 1.04 (95% CI: 1.01-1.07) and 1.03 (95% CI: 0.98-1.07) if the significant person was never hospitalized, 1.15 (95% CI: 1.08-1.23) and 1.24 (95% CI: 1.14-1.34) if the patient was hospitalized, 1.42 (95% CI: 1.27-1.57) and 1.45 (95% CI: 1.31-1.60) if admitted to the ICU, and 1.34 (95% CI: 1.22-1.46) and 1.36 (95% CI: 1.22-1.51) if the significant person died. Individuals of hospitalized, ICU admitted, or deceased patients showed higher prevalence of depression and anxiety during the entire 12 months after the COVID-19 diagnosis of the significant person.

**Conclusions:** Close friends and family members of critically ill COVID-19 patients show elevated prevalence of depression and anxiety throughout the first year after the diagnosis.

## Introduction

To date, over 0.6 billion people have been confirmed to have contracted the Coronavirus disease (COVID-19) at least once and approximately 6.5 million people have died due to COVID-19, across the globe (WHO, 12/10/2022). Given its worldwide influence, the substantial mental health impact of the pandemic has been increasingly documented. While most studies have focused on individuals directly exposed to the pandemic, especially patients with COVID-19^1,4^ and front-line healthcare workers^5,6^, families of individuals with COVID-19 might be another high-risk group for mental health problems and disorders^2^. Having a family member with SARS-CoV-2 infection has previously been associated with severe psychological distress^7^ and multiple studies have shown that having a relative suspected of or diagnosed with COVID-19 is associated with an increased risk of mental illness such as depression, anxiety, and stress-related disorders^1,2,8-11^.

Individuals who lost a family member due to COVID-19 might be the most vulnerable^2^. Bereavement has been consistently shown to increase the risk of mental illness^12,13^, including post-traumatic stress disorder (PTSD)^14^ and complicated grief^9,15^. Bereavement due to COVID-19 might be especially complicated, because the affected have limited possibilities to bid farewell to their loved ones, gather for mourning ceremonies, or receive support in their grief^9^. Verdery and colleagues created a prediction model indicating that every COVID-19-related death would leave approximately nine bereaved^16^. Given the accumulated number of deaths due to COVID-19 to date, this would mean up to 60 million COVID-19 bereaved individuals globally.

Starting in spring 2020, we leveraged five prospective cohort studies across four countries (Iceland, Norway, Sweden, and the UK) within the COVIDMENT consortium^17^, with the aim to examine the prevalence of symptoms of depression and anxiety among people whose family members or close friends (hereafter referred to as ‘significant persons’) contracted COVID-19.

We studied close friends, in addition to family members, as little data exists currently on how having a close friend with COVID-19 affects mental health. We focused on analyzing symptoms of depression and anxiety by the severity of COVID-19 in the significant person, hypothesizing a dose-response relationship between severity of COVID-19 and prevalence of symptoms.

## Methods

### Study design

The COVIDMENT network includes multiple cohort studies^17^. Since March 2020, all cohorts have been collecting self-reported data on COVID-19, including physical and mental health measures, using semi-harmonized questionnaires^17^. The following cohorts included questions on significant persons: the Icelandic COVID-19 National Resilience Cohort (C-19 Resilience), the Norwegian COVID-19 Mental Health and Adherence Study (MAP-19), the Norwegian Mother, Father, and Child Cohort Study (MoBa), the Swedish Omtanke2020 Study, and the UK-based CovidLife Study. We analyzed data collected from March 2020 to March 2022 in each of the five cohorts and performed a meta-analysis of the cohort-specific results to assess the association between having a significant person with COVID-19 and prevalence of symptoms of depression and anxiety. Data were collected once in MAP-19 (winter 2021/2022), twice in MoBa (summer 2020), thrice in C-19 Resilience (spring/summer 2020, winter 2020/2021, and summer 2021) and CovidLife (spring 2020, summer 2020, and winter 2020/2021), and up to 13 times in Omtanke2020 (monthly from June 2020 to February 2022), during the study period. Accordingly, we treated all variables related to COVID-19 as well as depression and anxiety as time varying.

We defined the cohort participants as exposed if they reported that a significant person had been diagnosed with COVID-19. The participants were accordingly classified as having “no significant person diagnosed”, “a significant person diagnosed before enrolment to the cohort”, or “a significant person diagnosed after enrolment”. There are also questions on hospitalization and admission to the ICU for COVID-19 in C-19 Resilience, MAP-19, and Omtanke2020 as well as death due to COVID-19 in MAP-19, Omtanke2020, and CovidLife. We accordingly classified the participants, whenever possible, as having “no significant person diagnosed”, “a significant person diagnosed but not hospitalized”, “a significant person diagnosed and hospitalized”, “a significant person diagnosed with ICU admission”, and “a significant person deceased due to COVID-19”. As one’s own diagnosis of COVID-19 might influence the mental health impact of having a significant person with COVID-19, we similarly ascertained the COVID-19 status for the cohort participants themselves.

We employed two validated instruments to measure symptoms of depression and anxiety, namely the 9-item Patient Health Questionnaire^18^ (PHQ-9) for depressive symptoms and the 7-item General Anxiety Disorder^19^ (GAD-7) for anxiety symptoms. On both instruments, a cut-off of ≥10 was used to define severe symptom load.^18-19^. We excluded participants who did not complete all items of the PHQ-9 and GAD-7 except in Omtanke2020 where, if at least 80% of the items were completed, we imputed the scores as described^20^. Our final analysis included 162,237 participants with data on depressive symptoms and 168,783 participants with data on anxiety symptoms. Among these participants, 24,718 (15.2%) and 27,003 (16.0%) reported having a significant person with COVID-19, respectively.

In addition to the COVID-19 status of the participants themselves, we included as covariables age at enrolment (i.e., baseline), sex or gender (male or female), educational level (“no formal education”, “compulsory, upper secondary, vocational, or other education”, “Bachelor’s/diploma university degree”, or “Master’s or PhD”), type of enrolment (by invitation or self-recruitment), relationship status (in a relationship or single), history of previous psychiatric disorders (yes or no), somatic comorbidities (no comorbidity, one comorbidity, two comorbidities, or >2 comorbidities), habitual drinking (yes or no, defined as ≥4 drinks consumed in one sitting for women or ≥5 drinks for men), and calendar period of enrolment. Type of enrolment was considered because participants enrolled by invitation differ from participants self-enrolled^21^. Some of the cohorts recruited participants via invitation to previously existing studies, some recruited participants through self-enrolment, whereas others used both. Further, as body mass index (BMI)^22-23^ and smoking^23-24^ have been associated with COVID-19 and mental health, we also included BMI (<25, 25-30, or >30 kg/m^2^) and smoking (no smoker, former smoker, or current smoker) as two additional covariables. We handled the missingness in categorical covariables using a separate category “missing”.

This study was approved by national or regional ethics review committees in Iceland (NBC no. 20–073, 21–071), Norway (REK 14140 and 125510), Sweden (DNR 2020-01785) and the UK (20/ES/0021). All participants provided written informed consent.

### Statistical analysis

First, to assess the impact of pandemic burden on the risk of depression and anxiety, we estimated the prevalence of depressive and anxiety symptoms among the cohort participants during the different calendar weeks of the study period when the respective cohort had an ongoing data collection against the burden of pandemic in the population (i.e., incidence of COVID-19 during the preceding two weeks of a specific calendar week of the corresponding country). The incidence of COVID-19 was obtained from government agencies^25-28^. We calculated the weekly prevalence among participants with or without a significant person with COVID-19 separately with marginal means using the EMMEANS R package^32^ and fitted a temporal trend of the prevalence estimates using a local regression (LOESS) model^33^.

Second, to assess the association between having a significant person with COVID-19 and risk of depression and anxiety, we calculated the prevalence ratio (PR) with 95% confidence interval (CI) of depression and anxiety, using the robust (modified) Poisson model with adjustment for intra-individual correlation for repeated measurements. In Model 1, we adjusted for age, sex or gender, COVID-19 status of the participants, and time of data collection (except for MAP-19 where data was collected only once). In Model 2, we additionally adjusted for educational level, type of recruitment, marital status, history of previous psychiatric disorders, somatic comorbidities, habitual drinking, BMI, and smoking. We calculated PR in relation to having a significant person with COVID-19 as well as by COVID-19 severity of the significant person. We first performed the analyses in each cohort and then performed a random-effects model meta-analysis of the aggregated data from each cohort to estimate the overall PR, using the R package METAFOR^34^. We used I² statistic to measure the heterogeneity between cohorts.

Finally, to understand the temporal relationship between having a significant person with COVID-19 and risk of depression and anxiety, we performed a separate analysis in Omtanke2020 with 13 monthly data collections. In this analysis, we defined time 0 as the month when the significant person was diagnosed. Among participants not reporting a significant person with COVID-19, we selected time 0 randomly to imitate that COVID-19 could have occurred any time during the study. For both groups, we calculated the prevalence of depression and anxiety up to 12 months before and up to 12 months after time 0. Among participants reporting a significant person with COVID-19, we calculated the prevalence by whether COVID-19 was diagnosed before or after enrolment as well as by disease severity (i.e., diagnosed but not hospitalized, hospitalized, ICU admitted, or deceased). The calculations were adjusted for age, sex or gender, COVID-19 status of the participant, time of data collection, and type of recruitment. The temporal trend of prevalence estimates was also smoothed using LOESS model.

## Results

**Table 1** shows the characteristics of the study participants. The mean age at enrolment ranged from 40.0 (MAP-19) to 56.9 (CovidLife), and proportion of female ranged from 60.3% (MoBa) to 81.5% (Omtanke2020). The percentage of reporting a significant person with COVID-19 ranged from 15.0% (C-19 Resilience) to 55.2% (Omtanke2020). Participants reporting a significant person with COVID-19 showed slightly higher (bi-)weekly prevalence of depression (top) and anxiety (bottom), compared with participants not reporting (**Figure 1a**). Although the result pattern was strongly related to the burden of pandemic in the population, the difference became stronger when analyzing the severity of COVID-19 in the significant person (**Figures 1b and 1c**).

**Table 1.** Descriptive characteristics of the five cohorts.

|  | Iceland<br>C-19 Resilience | Norway |  |  | Sweden | UK |
| --- | --- | --- | --- | --- | --- | --- |
|  |  | MoBa |  | MAP-19 | Omtanke2020 | CovidLife |
|  |  | Depression | Anxiety |  |  |  |
| Characteristics | Total<br>n (%) | Total<br>n (%) | Total<br>n (%) | Total<br>n (%) | Total<br>n (%) | Total<br>n (%) |
| Total (N) | 22898 | 91950 | 98496 | 2898 | 27752 | 16739 |
| Gender |  |  |  |  |  |  |
| Female | 16006 (69.9%) | 55411 (60.3%) | 58795 (59.7%) | 2326 (80.3%) | 22614 (81.5%) | 11172 (66.7%) |
| Male | 6892 (30.1%) | 36539 (39.7%) | 39701 (40.3%) | 572 (19.7%) | 5138 (18.5%) | 5567 (33.3%) |
| Age |  |  |  |  |  |  |
| Mean age (SD) | 54.4 (14.3) | 47.0 (5.2) | 47.0 (5.3) | 40.0 (14.0) | 48.7 (15.7) | 56.9 (14.1) |
| Median (IQR) | 56.0 [20.0] | 47.0 [46.0] | 47.0 [46.0] | 37.0 [21.0] | 49.0 [25.0] | 59.0 [19.0] |
| 18-29 years | 1487 (6.5%) | 0 (0.0%) | 0 (0.0%) | 837 (28.9%) | 3809 (13.7%) | 805 (4.8%) |
| 30-39 years | 2261 (9.9%) | 5944 (6.5%) | 6742 (6.8%) | 766 (26.4%) | 5069 (18.3%) | 1551 (9.3%) |
| 40-49 years | 4075 (17.8%) | 56000 (60.9%) | 60002 (60.9%) | 565 (19.5%) | 5308 (19.1%) | 2298 (13.7%) |
| 50-59 years | 5879 (25.7%) | 25578 (27.8%) | 26855 (27.3%) | 407 (14.0%) | 5961 (21.5%) | 3725 (22.2%) |
| 60-69 years | 5885 (25.7%) | 1209 (1.3%) | 1285 (1.3%) | 237 (8.2%) | 4417 (15.9%) | 5203 (31.1%) |
| 70 years + | 3311 (14.4%) | 57 (0.1%) | 67 (0.1%) | 86 (3.0%) | 3188 (11.5%) | 3157 (18.9%) |
| Missing | - | 3162 (3.4%) | 3545 (3.6%) | - | - | - |
| Education |  |  |  |  |  |  |
| Compulsory | 3297 (14.4%) | 1673 (1.8%) | 1894 (1.9%) | 131 (4.5%) | - | 1401 (8.4%) |
| Upper secondary, vocational, or other | 7095 (31.0%) | 25849 (28.1%) | 28249 (28.7%) | 1001 (34.5%) | - | 5741 (34.3%) |
| Bachelor's/diploma university degree | 7179 (31.3%) | 32248 (35.1%) | 34169 (34.7%) | 1766 (60.9%) | - | 3973 (23.7%) |
| Master's or Ph.D. | 5167 (22.6%) | 26816 (29.2%) | 28271 (28.7%) | - | - | 4298 (25.7%) |
| No formal education | - | 0 (0.0%) | 0 (0.0%) | - | - | 338 (2.0%) |
| Missing | 160 (0.7%) | 5364 (5.8%) | 5913 (6.0%) | - | 27752 (100%) | 988 (5.9%) |
| Marital status |  |  |  |  |  |  |
| In a relationship | 17522 (76.5%) | - | - | 1908 (65.8%) | 20106 (72.4%) | 12934 (77.3%) |
| Single | 5276 (23.1%) | - | - | 990 (34.2%) | 7508 (27.1%) | 3799 (22.7%) |
| Missing | 100 (0.4%) | - | - | - | 138 (0.5%) | 6 (<0.1%) |
| BMI (kg/m^2) |  |  |  |  |  |  |
| < 25, Normal or low weight | 6629 (28.9%) | 30066 (32.7%) | 30812 (31.2%) | 1014 (35.0%) | 14395 (51.9%) | 6534 (39.0%) |
| 25-30, Overweight | 8799 (38.4%) | 25530 (27.8%) | 26171 (26.6%) | 1012 (34.9%) | 8201 (29.5%) | 5895 (35.2%) |
| > 30, Obese | 6884 (30.1%) | 11697 (12.7%) | 11990 (12.2%) | 509 (17.6%) | 3769 (13.6%) | 4215 (25.2%) |
| Missing | 586 (2.6%) | 24657 (26.8%) | 29523 (30.0%) | 363 (12.5%) | 1387 (5.0%) | 95 (0.6%) |
| Current smoking |  |  |  |  |  |  |
| No, never | 10462 (45.7%) | 80338 (87.3%) | 79123 (80.3%) | - | 14297 (51.5%) | 10236 (61.2%) |
| No, former smoker | 8859 (38.7%) | - | - | - | 8459 (30.5%) | 5122 (30.6%) |
| Yes, currently | 3377 (14.7%) | 8417 (9.2%) | 8133 (8.3%) | - | 4645 (16.7%) | 1178 (7.0%) |
| Missing | 200 (0.9%) | 3195 (3.5%) | 11240 (11.4%) | - | 351 (1.3%) | 203 (1.2%) |
| Habitual drinking |  |  |  |  |  |  |
| Yes | 5140 (22.5%) | - | - | - | 7269 (26.2%) | 13126 (78.4%) |
| No | 17519 (76.5%) | - | - | - | 14950 (53.9%) | 3412 (20.4%) |
| Missing | 239 (1.0%) | 91950 (100%) | 98496 (100%) | - | 5533 (19.9%) | 201 (1.2%) |
| History of psychiatric disorders |  |  |  |  |  |  |
| Yes | 6501 (28.4%) | 14563 (15.8%) | 15642 (15.9%) | 731 (25.2%) | 9440 (34.0%) | 5386 (32.2%) |
| No | 16064 (70.2%) | 74135 (80.7%) | 79213 (80.4%) | 2167 (74.8%) | 17770 (64.0%) | 11251 (67.2%) |
| Missing | 333 (1.4%) | 3252 (3.5%) | 3641 (3.7%) | - | 542 (2.0%) | 102 (0.6%) |
| Somatic comorbidities |  |  |  |  |  |  |
| No comorbidity | 13419 (58.6%) | 73323 (79.8%) | 78358 (79.6%) | 2054 (70.8%) | 18144 (65.4%) | 9018 (53.9%) |
| One comorbidity | 6561 (28.7%) | 13543 (14.7%) | 14540 (14.7%) | 844 (29.2%) | 6227 (22.4%) | 4826 (28.8%) |
| Two comorbidities | 2107 (9.2%) | 1680 (1.8%) | 1797 (1.8%) | - | 1649 (5.9%) | 1909 (11.4%) |
| > Two comorbidities | 642 (2.8%) | 241 (0.3%) | 255 (0.3%) | - | 554 (2.0%) | 944 (5.6%) |
| Missing | 169 (0.7%) | 3163 (3.4%) | 3546 (3.6%) | - | 1178 (4.3%) | 42 (0.3%) |
| **Response period (baseline)** |  |  |  |  |  |  |
| April-June 2020 | 21448 (93.7%) | 91950 (100%) | 98496 (100%) | - | 1441 (5.2%) | 16739 (100%) |
| July-September 2020 | 261 (1.1%) | - | - | - | 10215 (36.8%) | - |
| October-December 2020 | 511 (2.2%) | - | - | - | 10768 (38.8%) | - |
| January-March 2021 | 636 (2.8%) | - | - | - | 1966 (7.1%) | - |
| April-June 2021 | 42 (0.2%) | - | - | - | 3357 (12.1%) | - |
| July-September 2021 | - | - | - | - | 5 (<0.1%) | - |
| October-December 2021 | - | - | - | - | - | - |
| January-March 2022 | - | - | - | 2898 (100%) | - | - |
| **Recruitment type** |  |  |  |  |  |  |
| Social media | - | - | - | 2898 (100%) | 11359 (40.9%) | - |
| Personal invitation from other cohort | - | 91950 (100%) | 98496 (100%) | - | 12057 (43.5%) | 4609 (27.5%) |
| Missing | - | - | - | - | 4336 (15.6%) | 12130 (72.5%) |
| **Participant's COVID-19 status** |  |  |  |  |  |  |
| Infected before baseline | 1017 (4.4%) | 198 (0.2%) | 194 (0.2%) | - | 1435 (5.2%) | 1683 (10.0%) |
| Infected during study | 183 (0.8%) | 9 (<0.1%) | 948 (1.0%) | 251 (8.7%) | 12285 (44.2%) | 796 (4.8%) |
| Not infected | 21653 (94.6%) | 91743 (99.8%) | 97354 (98.8%) | 2647 (91.3%) | 14032 (50.5%) | 14146 (84.5%) |
| Missing | - | - | - | - | - | 114 (0.7%) |
| **Participant's COVID-19 severity** |  |  |  |  |  |  |
| Not infected | 21881 (95.6%) | 91743 (99.8%) | 97354 (98.9%) | - | 14032 (50.5%) | 14146 (84.5%) |
| Infected but not hospitalized | 941 (4.1%) | 184 (0.2%) | 212 (0.2%) | - | 13594 (49.0%) | 2523 (15.1%) |
| Infected and hospitalized | 51 (0.2%) | 7 (<0.1%) | 8 (<0.1%) | - | 101 (0.4%) | 64 (0.4%) |
| Infected and with ICU admission | 25 (0.1%) | - | - | - | 25 (0.1%) | 3 (<0.1%) |
| Missing | - | 16 (<0.1%) | 922 (0.9%) | - | - | 3 (<0.1%) |
| **SP's COVID-19 status and severity** |  |  |  |  |  |  |
| Not infected | 19452 (85.0%) | - | - | 1884 (65.0%) | 12436 (44.8%) | 15073 (90.0%) |
| Infected but not hospitalized | 2886 (12.6%) | - | - | 964 (33.3%) | 12683 (45.7%) | - |
| Infected and hospitalized | 299 (1.3%) | - | - | 23 (0.8%) | 1145 (4.1%) | - |
| Infected and with ICU admission | 261 (1.1%) | - | - | 12 (0.4%) | 610 (2.2%) | - |
| Deceased | - | - | - | 15 (0.5%) | 878 (3.2%) | - |
| Missing | - | - | - | - | - | 1666 (10.0%) |

**Figure 1a.**
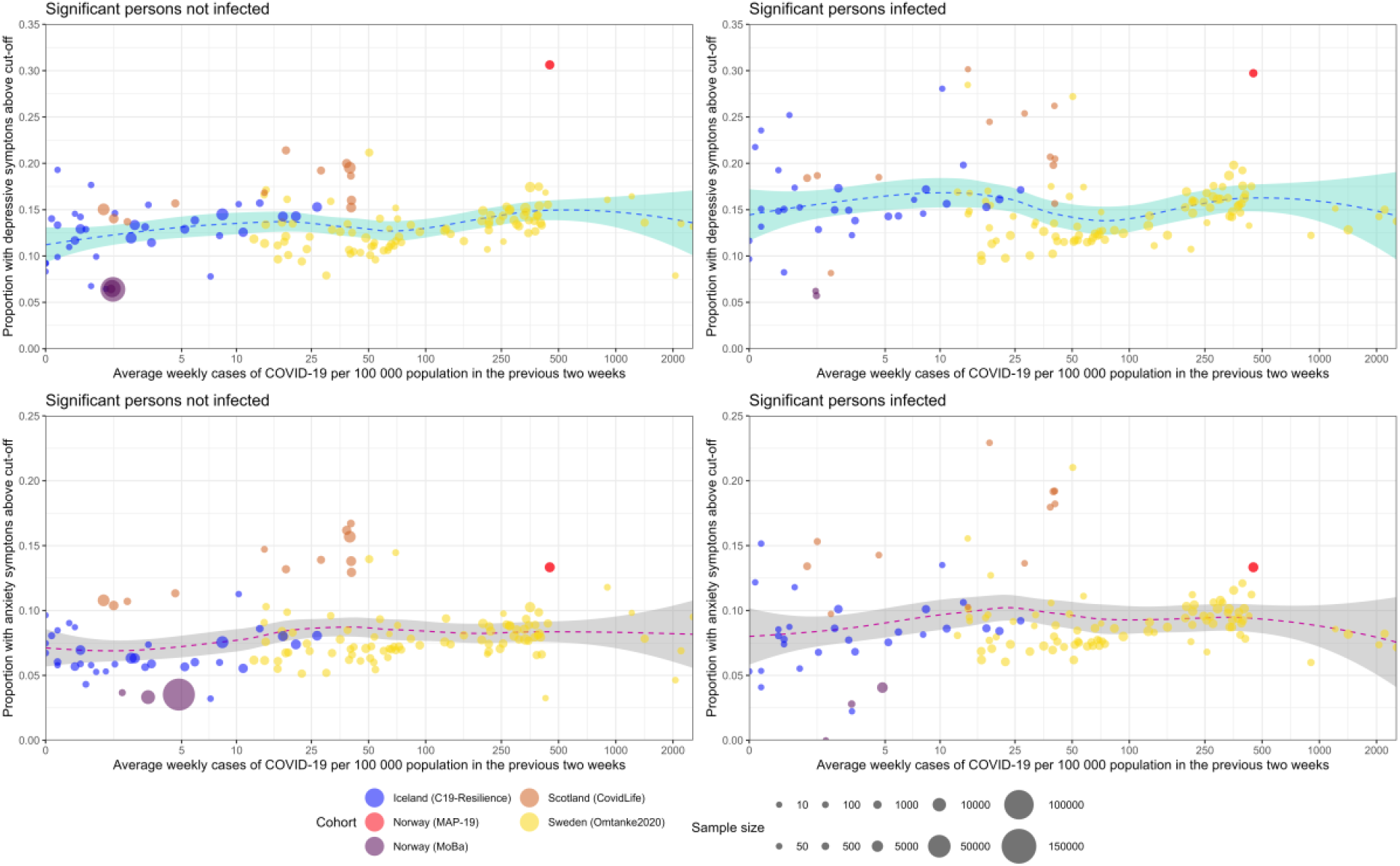
Depressive (top) and anxiety (bottom) symptoms and COVID-19 incidence across cohorts over the entire study period, stratified by infection of a significant person (no SP infected left, SP infected right). COVID-19 incidence is defined as the average number of confirmed cases per week per 100 000 persons in the 2 weeks prior to participant’s response to the PHQ-9 (depression) or GAD-7 (anxiety). Dotted blue line represents trend with 95% confidence interval (blue (depression)/grey (anxiety) area).

**Figure 1b.**
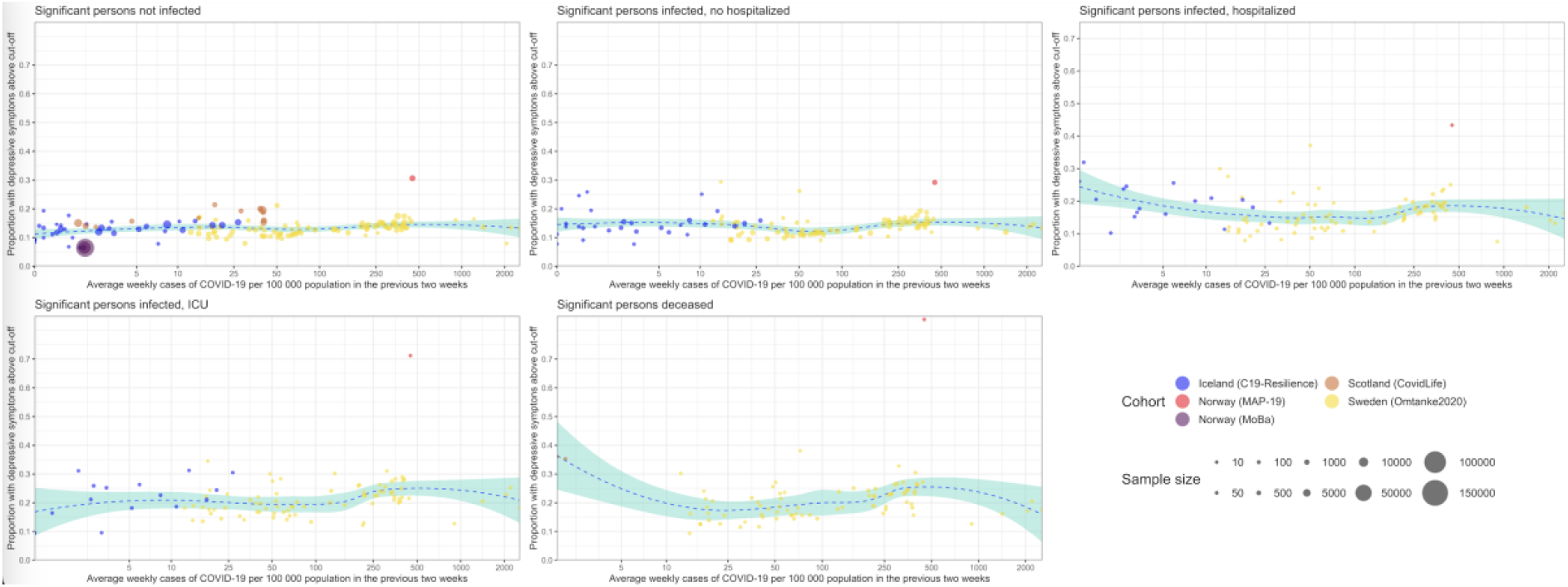
Depressive symptoms and COVID-19 incidence across cohorts over the entire study period, stratified by infection of a significant person by disease severity of the significant person. COVID-19 incidence is defined as the average number of confirmed cases per week per 100 000 persons in the 2 weeks prior to participant’s response to the PHQ-9 (depression). Dotted blue line represents trend with 95% confidence interval (blue area).

**Figure 1c.**
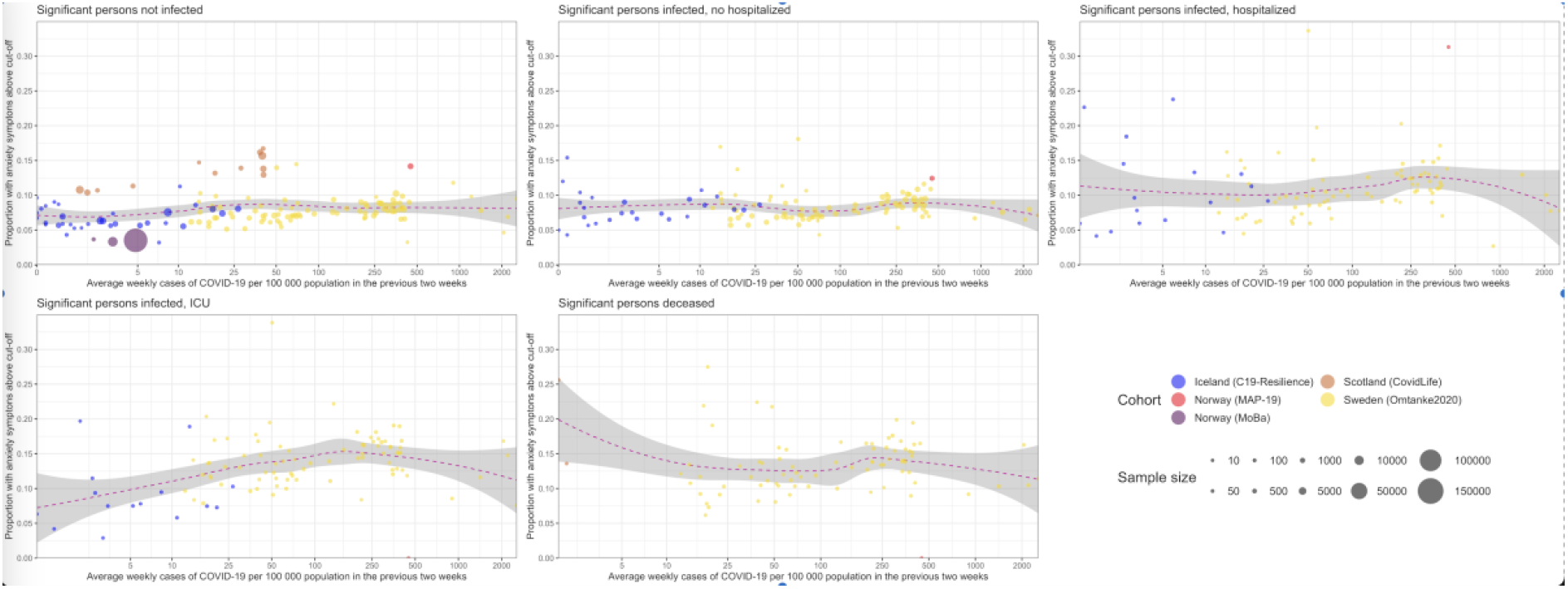
Anxiety symptoms and COVID-19 incidence across cohorts over the entire study period, stratified by infection of a significant person by disease severity of the significant person. The COVID-19 incidence is defined as the average number of confirmed cases per week per 100 000 persons in the 2 weeks prior to participant’s response to the GAD-7 (anxiety). Dotted blue line represents trend with 95% confidence interval (grey area).

Figure 2. shows the cohort-specific and pooled PRs of depression (left) and anxiety (right) in relation to having a significant person with COVID-19. In both Models 1 and 2, we found a positive association between having a significant person with COVID-19 and a higher prevalence of depression and anxiety in the pooled analyses, although the association for depression was not statistically significant in Model 1. There was less heterogeneity in Model 2 than Model 1 (i.e., I^2^ = 70% for depression and 67% for anxiety in Model 1 and <1% for depression and 31% for anxiety in Model 2).

For this reason, we present analyses based on Model 2 in **Figure 3** to show the results by disease severity of the significant person. Results on Model 1 can be found in **Supplementary Figure 1**. Apart from the analysis of anxiety in MAP-19, a partial dose-response relationship was noted in both the cohort-specific analyses and the pooled analyses, namely that the associations were strongest for having a significant person admitted to the ICU, followed by having a significant person hospitalized, for COVID-19. There was however no clear difference between having a significant person admitted to the ICU for COVID-19 and having a significant person deceased due to COVID-19. Excluding individual cohorts from the analyses did not change the results substantially. **Supplementary Figure 2** shows for example the results after excluding MoBa, the cohort with the largest sample size and youngest participants.

**Figure 2.**
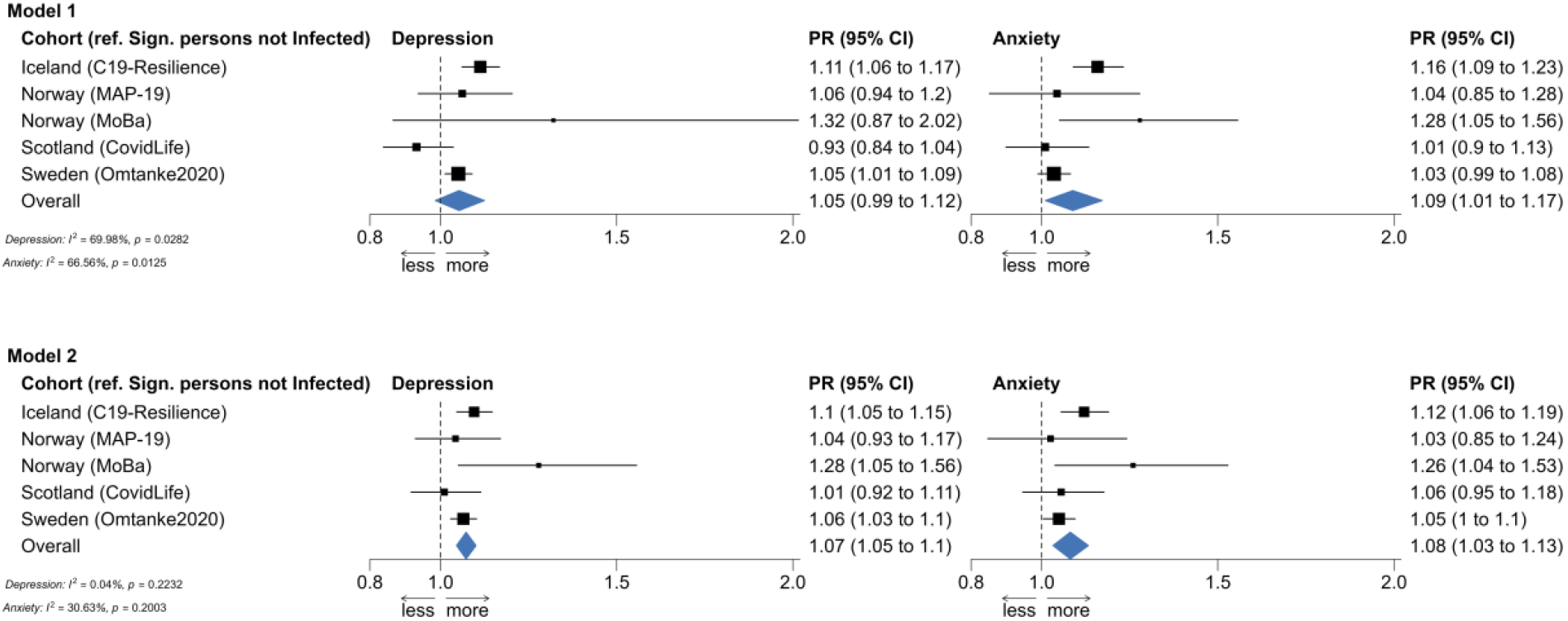
Prevalence ratios of depression (left) and anxiety (right) for those with and without (reference group) a significant person infected with SARS-CoV-2.

**Figure 3.**
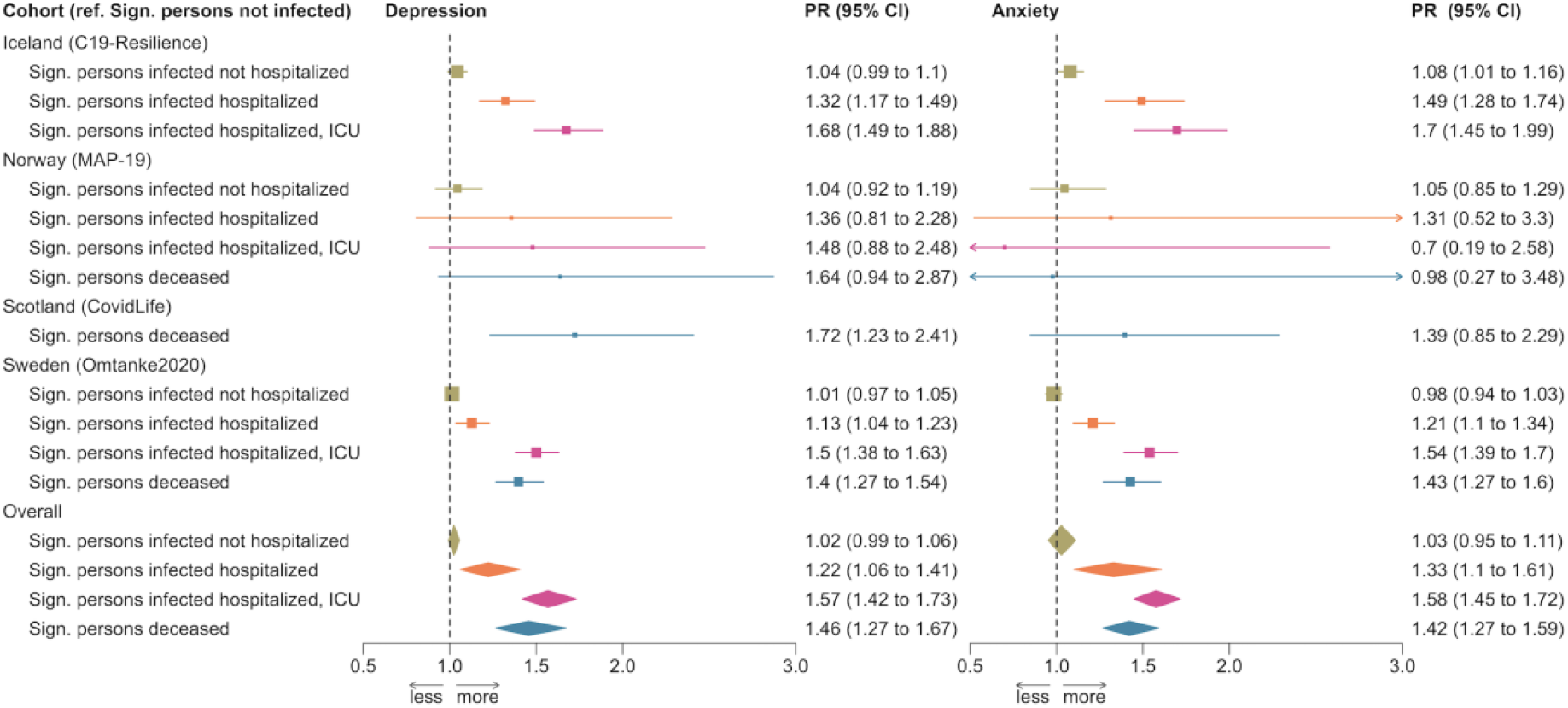
Prevalence ratios of depression (left) and anxiety (right) per disease severity of significant person after adjustment of all covariables.

In the Omtanke2020 cohort, the prevalence of depression and anxiety was much higher among participants reporting a significant person diagnosed with COVID-19 before enrolment, compared to participants not reporting such (**Figure 4**, top). Furthermore, the prevalence of depression and anxiety continuously decreased over time among participants not reporting a significant person with COVID-19, as well as among participants reporting a significant person diagnosed with COVID-19 before enrolment. The prevalence slightly increased around diagnosis of COVID-19 among participants reporting a significant person diagnosed with COVID-19 after enrolment. Participants reporting a significant person with COVID-19 without hospitalization demonstrated comparable prevalence of depression and anxiety as those not reporting a significant person with COVID-19 (**Figure 4**, bottom). However, those reporting a significant person hospitalized or admitted to the ICU for COVID-19, or deceased due to COVID-19, showed higher prevalence for both outcomes during the entire 12 months after the diagnosis of the significant person than those not reporting a significant person with COVID-19 or reporting a significant person with COVID-19 without hospitalization.

**Figure 4.**
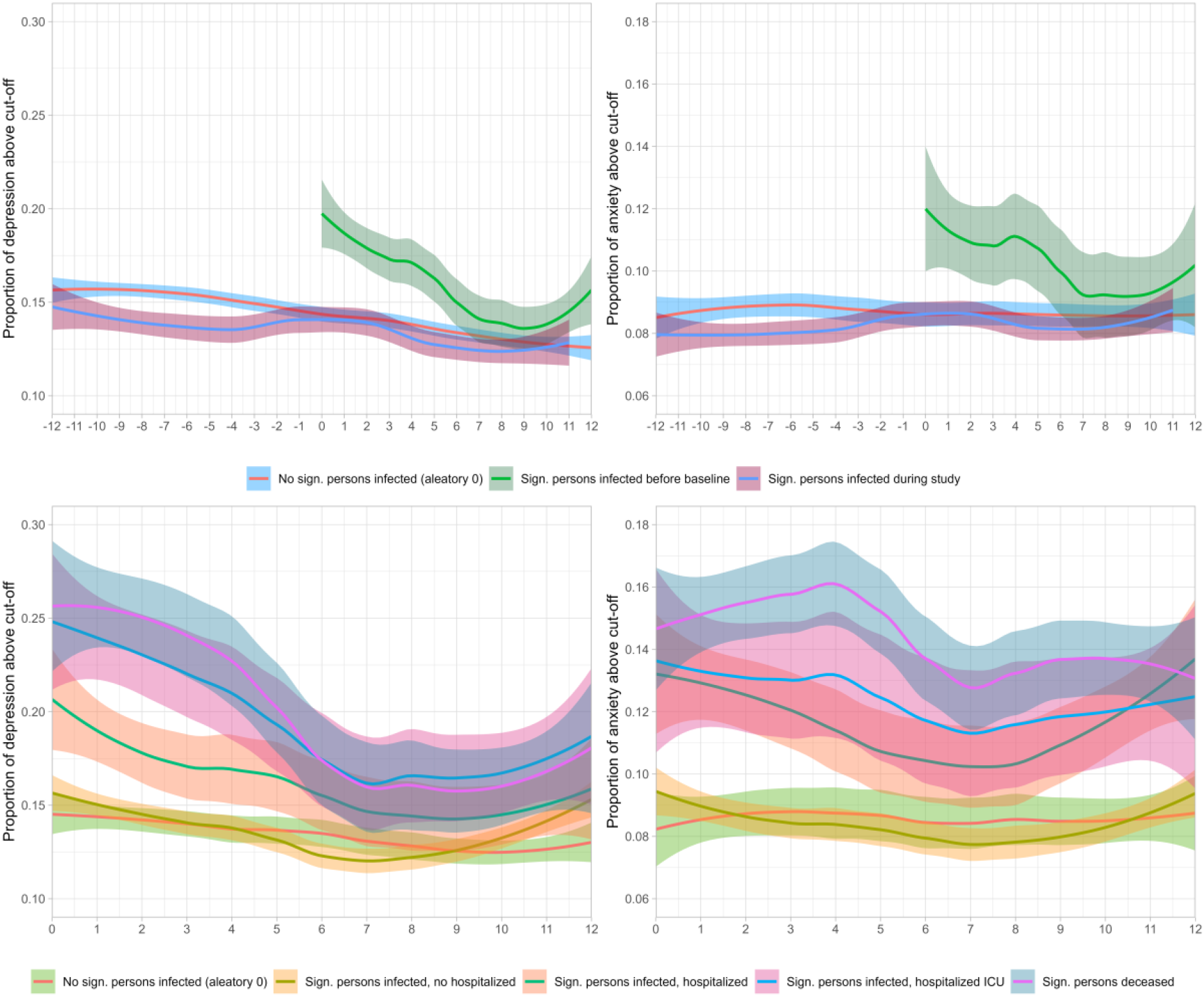
Time trends of monthly changes in depressive (left) and anxiety (right) symptoms for participants with/without SP infected (top), and by SP’s disease severity (bottom) in the Swedish Omtanke2020 cohort.

The difference was greatest immediately after COVID-19 diagnosis and decreased with time since diagnosis. Finally, there was a lower prevalence of depression among participants reporting a significant person hospitalized for COVID-19, compared with those reporting a significant person admitted to the ICU or deceased due to COVID-19. No clear pattern was, however, noted for anxiety.

## Discussion

In a study of over 160,000 individuals from four countries in Northern Europe, we found an elevated prevalence of depression and anxiety among individuals reporting having had a significant person (i.e., family members or close friends) diagnosed with COVID-19, particularly in cases of a critical COVID-19 illness (hospitalization, ICU, or death). This result was observed in the analysis of all individual cohorts as well as in the pooled analysis of all cohorts with data from the first 22 months of the pandemic.

Our finding of a higher prevalence of depression and anxiety among individuals reporting a significant person with COVID-19 is supported by few existing studies. One study showed a considerable risk of depression, anxiety, insomnia, and acute stress among family members and friends of patients with COVID-19 during the first wave of the pandemic in China^1^. Another study showed a prevalence of depression as 15% and of anxiety 16.3% among 153 relatives of COVID-19 patients^35^. A third study showed similarly prominent levels of depressive and anxiety symptoms between isolated COVID-19 patients and their relatives^8^. Our study is, to our knowledge, the first to demonstrate that the prevalence increment in mental health symptomology is proportional to the severity of COVID-19, mainly attributable to severe illness requiring inpatient or ICU care or leading to death.

We observed that individuals reporting a significant person hospitalized, admitted to the ICU, or deceased due to COVID-19 had persistently increased risk of depression and anxiety during the first year after the diagnosis of the significant person. Family members of COVID-19 patients admitted to the ICU have been shown to exhibit a higher prevalence and a slower improvement of depressive symptoms, compared to the patients themselves^36^. Half of the family members reported high levels of depressive symptoms one year later if the patient had used prolonged mechanical ventilation^36^. Further, symptoms of depression and anxiety did not seem to disappear even when the patients survived after ICU care^37^. Finally, family members have been reported to demonstrate high risk of PTSD^14^ and complicated grief^15^ following death of the patient in the ICU. Indeed, a higher level of prolonged grief disorder has been reported among individuals bereaved due to COVID-19^9^, regardless of ICU admission. These findings could be attributed to multiple factors, including fast transmission of the disease (no time to prepare), feelings of guilt (e.g., of perhaps having spread the illness to the patient), emotional shock of not being able to care or take farewell, fear of stigmatization^10^. Regardless, because of the extraordinarily large number of individuals deceased and the vast number of bereaved ones they left behind, bereavement due to COVID-19 has a substantial public health impact that will carry on for a long time to come^16^. Continued follow-up and surveillance is therefore needed for this risk population worldwide.

Strengths of our study include the large sample size, the long study period covering almost two years of the pandemic, the use of validated measures for depression and anxiety, and the availability of longitudinal data. Another distinct strength of the study is the cross-country design with harmonized or semi-harmonized data collection, leading to the unique opportunity of cross-validating findings between countries. A limitation of the study is the self-reported data on COVID-19 as well as depression and anxiety. However, potential measurement errors due to self-report would need to be systematic between reports on COVID-19 in a significant person and own depressive and anxiety symptoms, to explain the results of the study. The different definition of significant person between the cohorts is another limitation. Yet observing largely similar results across cohorts alleviates this concern to some extent. Finally, the study participants are all residing in European welfare states with relatively accessible health care for all, thus the findings cannot be readily generalizable to other populations.

In conclusion, significant persons of critically ill COVID-19 patients (i.e., who were hospitalized, required ICU admission, or died) show persistent elevations in symptoms of depression and anxiety. These findings motivate enhanced clinical surveillance of relatives and friends of patients suffering severe COVID-19 or other potential future pandemics.

## Data Availability

All cohort-level output data produced in the present study are available upon reasonable request to the authors. Individual-level raw data may be accessible after obtaining ethical approval from the respective ethical board due to data protection rules.

## Acknowledgements

This work was primarily supported with grants from Nordforsk (CovidMent, 105668) and Horizon2020 (CoMorMent, 847776). HA was supported by the Research Council of Norway (324620). Recruitment to CovidLife (UK) was facilitated by SHARE - the Scottish Health Research Register and Biobank. SHARE is supported by NHS Research Scotland, the Universities of Scotland and the Chief Scientist Office of the Scottish Government.

## Ethical approvals

The C-19 Resilience was approved by the National Bioethics Committee (NBC no. 20–073, 21–071) as well as the National Data Protection Authority. The establishment of MoBa was based on a license from the Norwegian Data Protection Agency and approval from The Regional Committees for Medical and Health Research Ethics (REK). The MoBa cohort is now based on regulations related to the Norwegian Health Registry Act. The current study was approved by REK (14140). MAP-19 was approved by REK (reference: 125510) and The Norwegian Centre for Research Data (reference: 802810). The Omtanke2020 Study was approved (DNR 2020-01785) by the Swedish Ethical Review Authority on 3 June 2020. The CovidLife study was reviewed and given a favourable opinion by the East of Scotland Research Ethics Committee (Reference: 20/ES/0021, AM02, AM04, AM05, AM11).

## Supplementary Tables and Figures

**Supplementary Figure 1.**
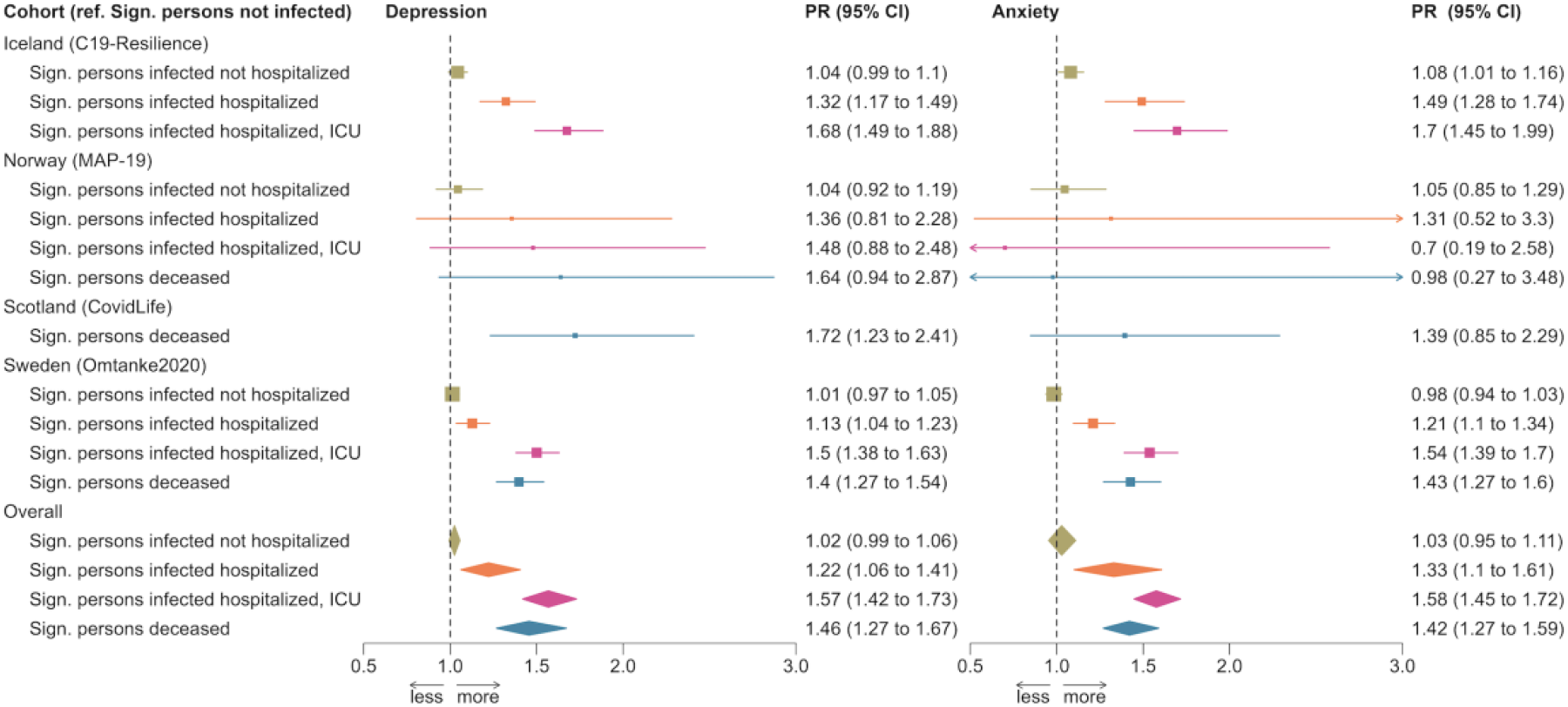
Prevalence ratios of depression (left) and anxiety (right) per disease severity of significant person after adjustment of all covariables. Model 1, adjusted for age, sex or gender, COVID-19 status of the participants, and time of data collection.

**Supplementary Figure 2.**
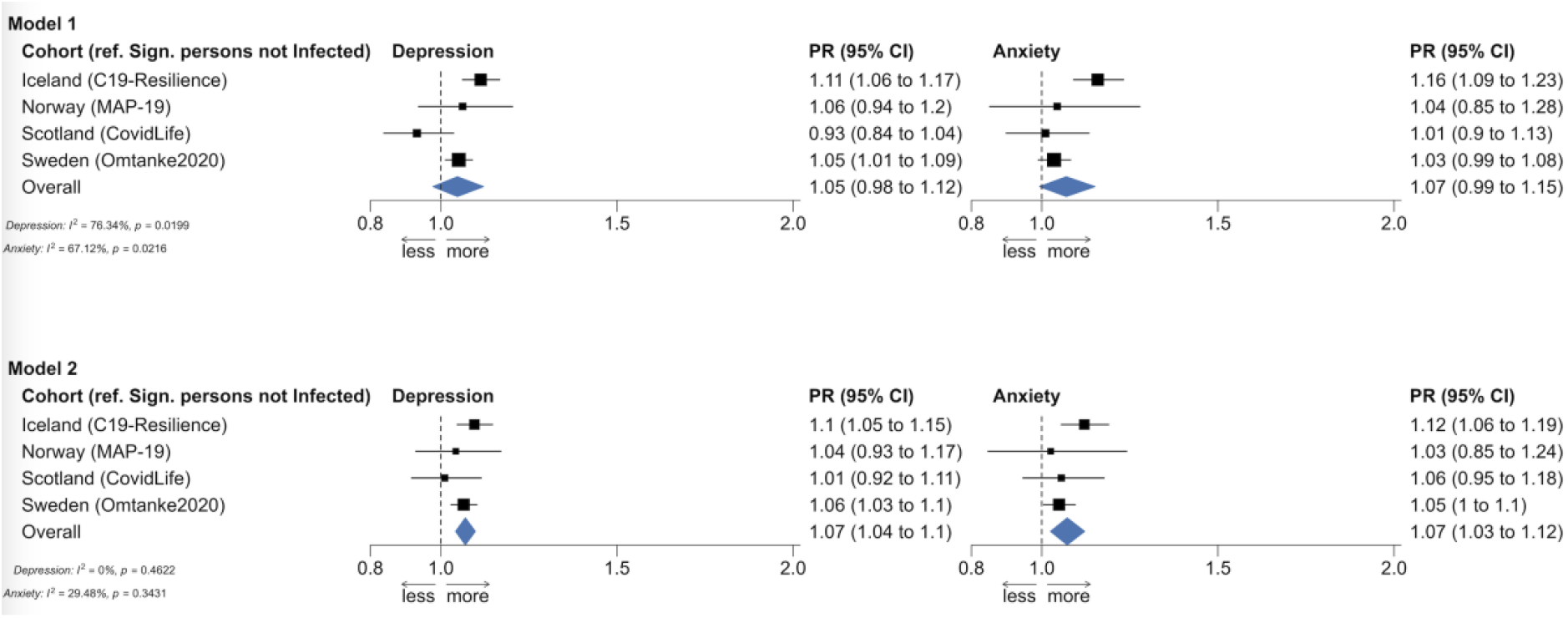
Figure 2 without the Norwegian MoBa cohort.

